# Association Between Telehealth Use and Downstream 30-Day Medicare Spending

**DOI:** 10.1101/2025.01.31.25321423

**Authors:** Chad Ellimoottil, Ashwin J. Kulkarni, Ziwei Zhu, Rodney L. Dunn, Chiang-Hua Chang, Elena Chun, Hechuan Hou, James D. Lee, Jeffrey S. McCullough, Michael P. Thompson

## Abstract

**Objectives:** The objective of this study was to investigate whether healthcare visits initiated by telehealth had higher or lower 30-day spending compared to in-person-initiated visits. The study compared the overall spending, rates of return visits, laboratory tests, and imaging procedures within 30 days for Medicare fee-for-service patients who underwent in-person and telehealth evaluations between July 1, 2020 and December 31, 2022.

**Study Design:** A large-scale retrospective cohort study using propensity score matching.

**Methods:** This study included 100% of Medicare fee-for-service beneficiaries who are aged >65 years while excluding those with Medicare Advantage coverage and those without continuous Medicare Parts A and B. We identified patients with no prior visits and 30-day episodes of care initiated by outpatient telehealth or in-person visits from July 1, 2020 through December 31, 2022. We then compared adjusted 30-day total Medicare spending, rates of return visits, laboratory tests, and imaging utilization between propensity-matched index visits initiated by telehealth versus in-person.

**Results:** Telehealth-initiated visits were associated with lower 30-day spending ($260 vs. $342; net: −$82), though return visit rates were higher for telehealth (16.1% vs. 14.1%). Both lab test rates (7.8% vs. 24.2%) and imaging rates (3.5% vs 7.8%) were lower for telehealth-initiated episodes compared to in-person-initiated episodes.

**Conclusions:** Telehealth-initiated episodes of care were associated with lower 30-day Medicare spending and reduced utilization of labs and imaging. These findings suggest that telehealth, when used as a substitute for office visits, may reduce overall Medicare spending.

## INTRODUCTION

Since March 2020, telehealth has become an integral part of healthcare delivery.^1,2^ Among Medicare beneficiaries, the use of telehealth services surged in early 2020, followed by a gradual decline.^3^ In 2022, 43% of patients had at least one evaluation and management visit via telehealth.^4^ This expansion was facilitated by telehealth coverage flexibilities introduced during the national public health emergency. The Consolidated Appropriations Act, 2023, extended several key Medicare telehealth policies through December 31, 2024.^5^ The American Relief Act, 2025, further extended temporary telehealth coverage through March 31, 2025, as part of the continuing resolution.^6^ However, the extension of these flexibilities beyond that date remains uncertain, and the 119^th^ Congress may be prompted to consider permanent telehealth policies before the government funding agreement expires in March, 2025.

There has been bipartisan support for telehealth in the 118th Congress, for example, as evidenced by the House Ways and Means Committee’s unanimous passage of the Preserving Telehealth, Hospital, and Ambulance Access Act (H.R. 8261), which would extend many of Medicare telehealth flexibilities through 2026.^7^ However, despite the bipartisan enthusiasm for making temporary telehealth coverage policies permanent, the lack of data on the impact of telehealth on Medicare spending had hindered policymaking for a permanent legislation.

In fact, concerns over telehealth’s effect on Medicare expenditures are a primary reason why temporary extensions were granted instead of permanent policies. The Congressional Budget Office has raised several unresolved questions regarding telehealth, particularly whether these services substitute for in-person Medicare services or are provided in addition to them.^8^ A crucial question is whether the use of telehealth services influences beneficiaries’ likelihood of utilizing downstream services. On one hand, insufficient telehealth visits may lead to redundant expenses and potentially require additional diagnostic procedures. On the other hand, telehealth could reduce Medicare spending by enabling early intervention, thereby reducing the need for emergency room visits and hospital admissions. Furthermore, telehealth might lessen the frequency of unnecessary diagnostic tests often performed during physical appointments, addressing the issue of "convenience testing" without clear medical justification.

Several prior studies have investigated the effects of telehealth on healthcare expenditures. An analysis by the Medicare Payment Advisory Commission (MedPAC) found that hospital service areas (HSAs) with more intensive telehealth use experienced slightly higher overall healthcare costs per beneficiary.^9^ Additionally, Nakamoto et al. observed a $248 increase in per-patient-per-year spending associated with telemedicine use across health systems.^10^ While both studies employed the difference-in-difference (DiD) technique to control for secular trends and enhance causal inference, attributing the rise in costs directly to telehealth remains challenging due to the aggregated nature of the outcomes.^11^

Our approach to addressing this limitation is to conduct an episode-level study. By directly examining the use of return visits, labs, and imaging within 30 days of a telehealth visit compared to similar in-person visits, we aim to expand on the existing literature by focusing on the downstream costs that follow a telehealth encounter.

## METHODS

### Data Sources and Population

We utilized a dataset comprising all claims for 100% national fee-for-service Medicare beneficiaries. The study encompassed patients who received care between July 1, 2020, and December 31, 2022. By starting our study period on July 1, 2020, we aimed to minimize the influence of care disruptions that occurred early during the pandemic. To construct our dataset, we used the Medicare Master Beneficiary Summary and excluded beneficiaries who were less than 65 years old or above 99 years old, not continuously enrolled in both Medicare Part A and Part B, as well as those with Medicare Advantage coverage for the specified year. This study followed the STROBE reporting guidelines and was determined to be exempt from review by the University of Michigan Institutional Review Board.^12^

### Identifying Episodes of Care

Using the Carrier file, we identified outpatient evaluation and management (E&M) visits received by our study population, focusing on Berenson-Eggers Type of Service (BETOS) codes (M1A, M1B, M5B, M5C, M5D). To identify telehealth services, we took two steps. First, we looked for outpatient E&M services with the appropriate modifier codes (GT, GQ, 95) or place of service code (02). Secondly, we ensured that the Healthcare Common Procedure Coding System (HCPCS) codes associated with the identified claims were included in Medicare’s list of eligible telehealth services for the corresponding year or were classified by Medicare as phone services.

We then mapped each E&M visit to a diagnosis category using the Clinical Classifications Software Refined (CCSR, v2022.1) based on the primary diagnosis code. Developed as part of the Healthcare Cost and Utilization Project, CCSR aggregates 70,000 International Classification of Diseases, 10th Revision, Clinical Modification/Procedure Coding System (ICD-10-CM/PCS) codes into over 530 clinically meaningful diagnosis categories.

For defining an episode of care, we considered the 30-day window following an index outpatient visit. Index visits were identified as those with no other outpatient E&M visits in the same diagnosis category as the initial code within the preceding 60 days. In cases where we found multiple visits for the same primary diagnosis code on the same day, we prioritized telehealth visits first, followed by office visits for the index visit definition. A maximum of one episode was constructed for each beneficiary-CCSR combination every 60 days.

### Outcomes

Our primary outcome was 30-day total Medicare payment, defined as the total payments from the same CCSR paid by the Medicare program during the index visit and up to 29 days after the index visit. This outcome did not include coinsurance or payments made by the patient because our study is focused on Medicare spending. We examined total Part B payments, outpatient payments, and inpatient payments during the 30-day period.

In addition to cost, we also examined several utilization outcomes. First, we examined the 30-day related return visit rate, which we defined as the rate of episodes where there was a second office visit for the same CCSR within 29 days after the index visit day. Similarly, we examined the rate of episodes that included related imaging and labs during the index visit day or within 29 days after the day of the index visit. Imaging and lab tests were identified using BETOS categories (I and T, respectively). Imaging and lab tests were considered “related” if the diagnosis code category for primary diagnosis associated with the test was in the same CCSR as the index visit.

### Statistical Analysis

We performed an episode-level analysis where we compared our primary and secondary outcomes between episodes of care that were initiated by an in-person index visit versus a telehealth index visit. To account for the differences in the two groups, we performed propensity score matching. To do so, we first built a multivariable logistic regression model to predict whether the episode of care would be telehealth or in-person. In this model, we included the following predictors: CCSR, gender, age, race, rural, dual eligibility, and Hierarchical Condition Category Risk Adjustment Factor scores. Using this model, we obtained the predicted propensity that the episode would be a telehealth visit. We then performed nearest neighbor matching with replacement to find one control within each CCSR for each telehealth episode. After creating the two balanced cohorts, we performed a paired t-test for each continuous outcome and McNemar’s test for each categorical outcome to evaluate differences between telehealth and in-person episodes.

### Sensitivity Analysis

We performed several sensitivity analyses to assess the robustness of our results. First, we examined outcomes stratified by telehealth propensity quintiles and selected CCSR categories in mental health and non-mental health. We also assessed the outcomes using matching without replacement where each in person control episode is matched to a maximum of one telehealth episode.

## RESULTS

### Study Population

The study included 30,079,958 participants with 36,709,528 and 429,891,125 episodes initiated by telehealth and in-person visits, respectively. Mean age was 76 for telehealth group and 75 for in-person group, with the telehealth group having higher risk adjustment factor scores (1.29 vs. 0.83) correlating with increased average comorbidities, higher proportion of female patients (59% vs. 54%), and a lower proportion of rural patients (17% vs. 27%).

### 30-Day Spending

Telehealth-initiated visits were associated with lower 30-day spending ($260 vs. $342; difference, −$82 [95% CI: -83 to -82]) **(Table 1)**. This decreased spending was consistent across multiple cost categories. Telehealth was associated with lower 30-day inpatient spending ($59 vs. $71, -$11 [95% CI: -12 to -11]), lower 30-day outpatient spending ($56 vs. $76, -$20 [95% CI: -21 to -20]), and lower 30-day Medicare Part B spending ($145 vs. $196, -$51 [95% CI: -51 to -50]). Spending was consistently lower for telehealth across all propensity quintiles **(Appendix Table 1)**.

**Table 1.** 30-Day Spending Composition of In-Person vs. Telehealth-Initiated Care Episodes.

|  | **In-Person** | **Telehealth** | **Difference** | **95%**<br>**Confidence**<br>**Intervals for**<br>**Difference** | **P-Value** |
| --- | --- | --- | --- | --- | --- |
| **Inpatient 30-Day Spending** | \$71 | \$59 | -\$11 | (-\$12 to -\$11) | <0.001 |
| **Outpatient 30-Day Spending** | \$76 | \$56 | -\$20 | (-\$21 to -\$20) | <0.001 |
| **Part B 30-Day Spending** | \$196 | \$145 | -\$51 | (-\$51 to -\$50) | <0.001 |
| **Total 30-Day Spending** | \$342 | \$260 | -\$82 | (-\$83 to -\$82) | <0.001 |

### 30-Day Return Visit Rates

Rates of 30-day return visits for related conditions were higher for telehealth-initiated visits compared to in-person-initiated visits (16.1% vs. 14.1%, +2.0% [95% CI: 2.0 to 2.0]) **(Table 2)**. All quintiles show an increase in return visit rates for telehealth **(Appendix Table 2)**.

**Table 2.** 30-Day Return Visit, Imaging, and Lab Test Utilization Rates for In-Person vs. Telehealth-Initiated Episodes.

|  | **In-Person** | **Telehealth** | **Difference** | **95% Confidence**<br>**Intervals for**<br>**Difference** | **P Value** |
| --- | --- | --- | --- | --- | --- |
| **Return Visit Rate** | 14.1% | 16.1% | 2.0% | (2.0% to 2.0%) | <0.001 |
| **Image Rate** | 7.8% | 3.5% | -4.4% | (-4.4% to -4.3%) | <0.001 |
| **Lab Test Rate** | 24.2% | 7.8% | -16.4% | (-16.4% to -16.4%) | <0.001 |

### 30-Day Imaging and Lab Tests Rates

In contrast to return visit rates, rates of imaging tests completed within 30 days of the index visit were lower for telehealth-initiated visits compared to in-person-initiated visits (3.5% vs. 7.8%, - 4.4% [95% CI: -4.4 to -4.3] **(Table 2)**. Imaging rates were largely variable by propensity quintiles, though each quintile did show a significant decrease in imaging rates for telehealth visits **(Appendix Table 2)**. To an even greater degree, the rate of lab tests completed within 30 days was also lower for telehealth-initiated visits (7.8% vs. 24.2%, -16.4% [95% CI: -16.4 to - 16.4]. This pattern is consistent across propensity quintiles, with differences between telehealth and in person visits ranging from 8.9% to 23.7%. Overall, telehealth-initiated visits were associated with lower imaging and lab utilization.

### Mental Health CCSRs

The three largest mental health CCSR categories included depression-related disorders, anxiety and fear-related disorders, and trauma-and stressor-related disorders. In depression-related disorders (MBD002), telehealth-initiated visits were associated with lower 30-day spending ($220 vs. $271, −$51). However, telehealth-initiated visits were associated with higher 30-day spending in both anxiety and fear-related disorders (MBD005) ($167 vs. $146, +$20) and trauma and stressor-related disorders (MBD007) ($241 vs. $214, +$28). For all three categories, telehealth-initiated visits had higher return visit rates but lower imaging and lab test rates **(Appendix Table 3).**

### Non-Mental Health Conditions

The three largest non-mental health CCSR categories included essential hypertension, sleep-wake disorders, and both infective and non-infective spondyloarthropathies. In all of these categories, telehealth-initiated visits were associated with lower 30-day spending: essential hypertension (CIR007) ($112 vs. $126, -$14), sleep wake disorders (NVS016) ($107 vs. $155; - $47), and spondyloarthropathies (MUS011) ($416 vs. $568, -$152). For all three categories, telehealth-initiated visits once again had higher return visit rates but lower imaging and lab test rates **(Appendix Table 4).**

### Sensitivity Analysis

When performing a sensitivity analysis to assess the outcomes using matching without replacement, we found that our results were consistent. Specifically, telehealth-initiated visits were associated with lower spending, higher return visits, and lower imaging and lab tests **(Appendix Table 5).**

## DISCUSSION

In this analysis of 30-day Medicare spending for telehealth and in-person episodes of care, we found that episodes initiated via telehealth were associated with lower 30-day expenditures compared to those initiated in person. This reduction in spending was consistent across inpatient, outpatient, and Part B categories. Although telehealth episodes resulted in a higher frequency of 30-day follow-up visits, they were associated with fewer laboratory and imaging tests, likely contributing to the overall decrease in 30-day Medicare spending for telehealth-initiated care episodes.

Our finding that telehealth is associated with lower Medicare spending contrasts with the conclusions drawn by the MedPAC and Nakamoto et al.^9,10,13^ MedPAC analyzed the 6-month total cost of care for Part A and/or B services per fee-for-service Medicare beneficiary across HSAs with varying levels of telehealth use (low, medium, or high terciles). They found that HSAs with high telehealth use had a higher total cost of care, with an additional $164.99 per beneficiary over six months compared to areas with low telehealth use. MedPAC employed a DiD analysis to improve causal inference and adjust for secular trends.

Similarly, Nakamoto et al. used a DiD framework but applied it to a health system-level analysis. Their research revealed that Medicare patients receiving care in health systems within the highest quartile of telemedicine use experienced a $248 increase in per-patient, per-year spending. The primary advantage of the DiD approach is its ability to control for unobserved, time-invariant confounders. By comparing changes in outcomes over time between treatment groups (high telehealth tercile or quartile) and control groups (low telehealth use), the DiD method effectively removes the influence of factors that remain constant over time, isolating the impact of telehealth use on outcomes.

However, there are also limitations to this approach. First, unobserved differences between HSAs and health systems that evolved over time, especially during the pandemic, could have confounded the results, leading to biased estimates of the association between telehealth use and outcomes. This is particularly relevant given the substantial changes in healthcare delivery beyond telehealth during this period.^3^ Second, aggregating data at the HSA level may obscure important variations at the individual or provider level. Differences in telehealth use within HSAs or variations in how telehealth was implemented across health systems could lead to heterogeneous effects that the DiD approach might not capture, potentially resulting in an oversimplified interpretation of the results.

In response to these limitations, our study focused on a shorter interval—30-day care episodes— hypothesizing that a more confined time frame might better correlate downstream expenditures with the initial telehealth encounter. Additionally, we employed propensity score matching to enhance our dataset by accounting for inherent cost differences between telehealth and in-person-initiated episodes. However, like a DiD analysis, a propensity score-based episode analysis may still be subject to unmeasured confounding variables, underscoring the importance of utilizing diverse analytical strategies to inform policy decisions.

In addition to reduced Medicare spending, our study also found a significant decrease in both imaging and lab test rates within the telehealth cohort. While multiple prior studies have examined healthcare utilization, including lab tests and imaging, for audio versus video telehealth visits, none have directly compared these metrics with in-person care.^14,15^ Although it’s not entirely clear whether the reduced utilization of labs and imaging is the sole driver of decreased spending, this seems likely given the increased frequency of 30-day return visits following telehealth-initiated care episodes in our study. This finding is consistent with previous literature from our group and others. For example, a 2021 study on follow-up care for acute respiratory infections reported that 10.3% of telehealth consultations led to subsequent care, compared to 5.9% for in-person visits.^16^

### Limitations

Our study has several limitations. First, our study utilized Medicare Fee-for-Service data; hence, our results may not extend to patients with Medicare Advantage or other commercial plans. However, understanding telehealth’s impact on Medicare spending is crucial for Medicare policy decision-making. Second, despite employing matching to equilibrate our control and intervention groups, unmeasured confounders may still bias our findings. Third, while 30-day period after an E&M visit likely captures most downstream utilization related to the index visit, our findings may underestimate downstream spending as some follow-up care may take longer than 30 days to complete. Fourth, the scope of our analysis was confined to the costs and utilization associated with telehealth, without evaluating the quality of care between telehealth and in-person visits. Similarly, the differential downstream rates of return visits, lab tests, and imaging utilization we found may impact important clinical outcomes that are not evaluated in this study.

These limitations notwithstanding, our study offers critical insights for healthcare providers, payers, and, most importantly, policymakers engaged in ongoing debates regarding the permanent coverage of telehealth services. In the context of Medicare spending, understanding telehealth’s financial implications remains paramount. Our research finds that telehealth visits are not linked to increased 30-day subsequent healthcare costs. This finding is primarily attributed to the observed decrease in laboratory tests and imaging procedures. Such evidence is poised to inform policy discussions, underscoring the cost-effectiveness of telehealth in contemporary healthcare delivery models.

## CONCLUSIONS

In this three-year retrospective cohort study, we found that telehealth-initiated episodes of care were associated with lower 30-day Medicare spending. Additionally, the telehealth cohort exhibited lower rates of lab and imaging utilization, accompanied by higher return visit rates, trends observed across both mental health and non-mental health conditions. These findings suggest that telehealth may not contribute to increased downstream spending. This evidence is crucial for healthcare policymakers as they consider the financial implications of permanent telehealth legislation for Medicare.

**Figure 1.**
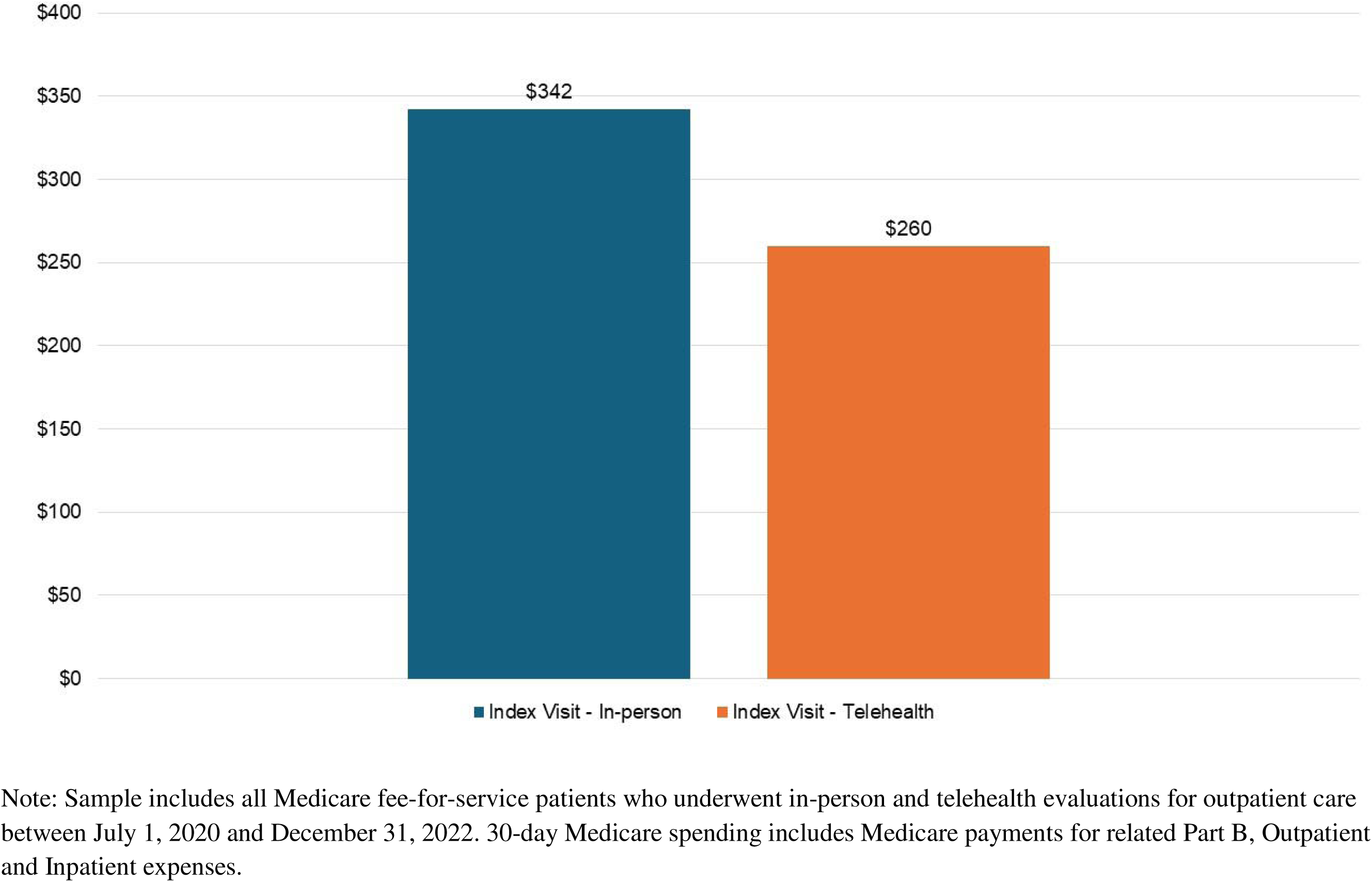
30-Day Medicare Spending of In-Person vs. Telehealth-Initiated Episodes

**Figure 2.**
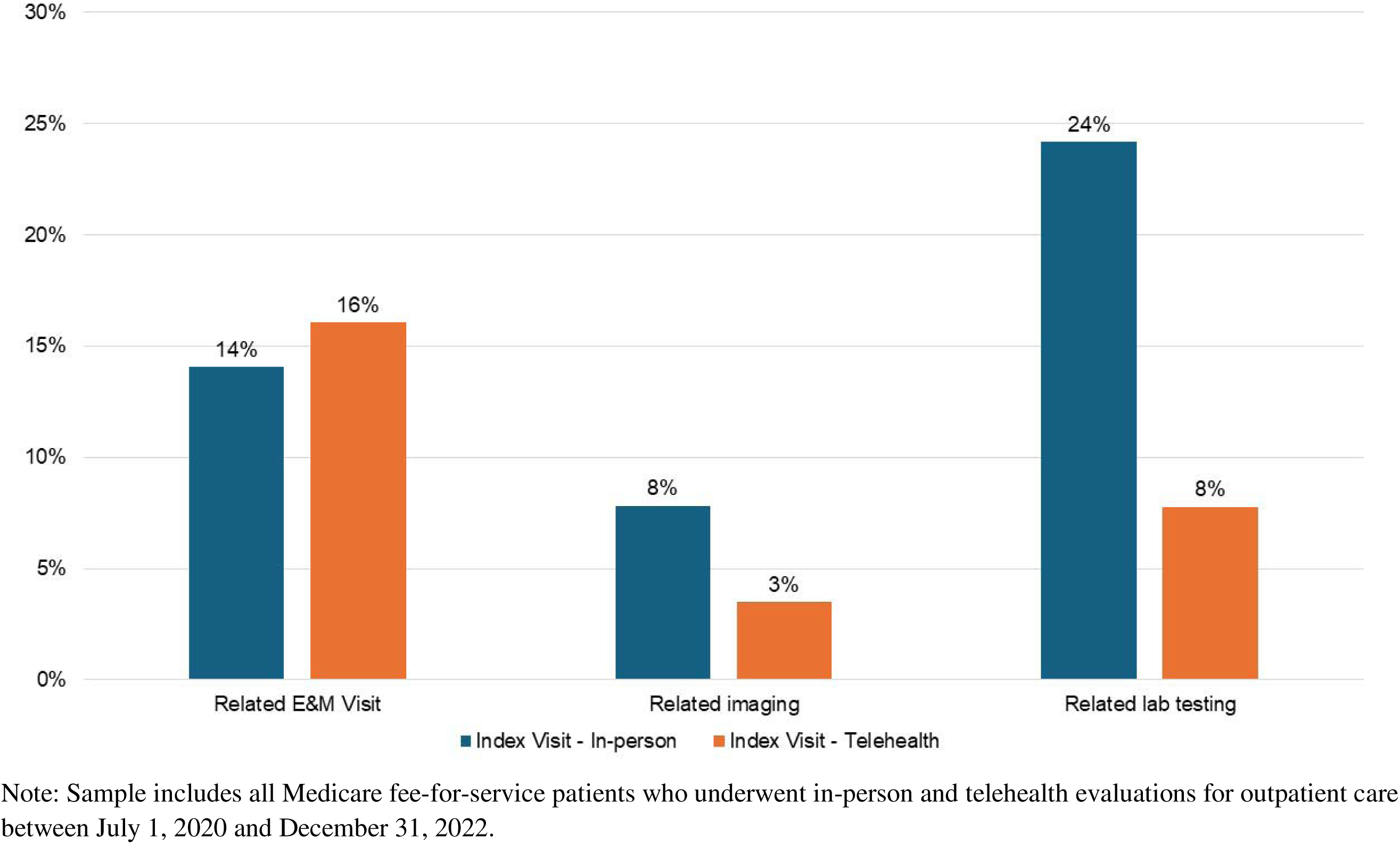
30-Day Return Visit, Imaging, and Lab Test Utilization Rates for In-Person vs. Telehealth-Initiated Episodes

## Data Availability

All data in the study are publicly available through the Chronic Conditions Data Warehouse (www2.ccwdata.org).

