## Appendix Tables for "Association Between Telehealth Use and Downstream 30-Day Medicare Spending"

**Appendix Table 1** – 30-Day Spending Composition of In-Person vs. Telehealth-Initiated Care Episodes by Propensity Quintiles

|  | **Inpatient Spending** | | | **Outpatient Spending** | | | **Part B Spending** | | | **Total Spending** | | |
| --- | --- | --- | --- | --- | --- | --- | --- | --- | --- | --- | --- | --- |
|  | **In-Person** | **Tele**  **health** | **Diff** | **In-Person** | **Tele**  **health** | **Diff** | **In-Person** | **Tele health** | **Diff** | **In-Person** | **Tele health** | **Diff** |
| **Quintile 1** | $13 | $21 | $8 | $54 | $73 | $19 | $176 | $126 | -$50 | $243 | $219 | -$24 |
| **Quintile 2** | $36 | $38 | $2 | $84 | $67 | -$17 | $192 | $137 | -$55 | $312 | $242 | -$70 |
| **Quintile 3** | $66 | $62 | -$4 | $92 | $74 | -$18 | $184 | $139 | -$45 | $342 | $275 | -$67 |
| **Quintile 4** | $62 | $53 | -$9 | $89 | $62 | -$27 | $215 | $149 | -$66 | $366 | $265 | -$102 |
| **Quintile 5** | $85 | $67 | -$18 | $64 | $43 | -$21 | $192 | $148 | -$44 | $340 | $258 | -$83 |

**Appendix Table 2** – 30-Day Return Visit, Imaging, and Lab Test Utilization Rates for In-Person vs. Telehealth-Initiated Episodes by Propensity Quintiles

|  | **Return Visit Rate** | | | **Imaging Rate** | | | **Lab Test Rate** | | |
| --- | --- | --- | --- | --- | --- | --- | --- | --- | --- |
|  | **In-Person** | **Telehealth** | **Diff** | **In-Person** | **Telehealth** | **Diff** | **In-Person** | **Telehealth** | **Diff** |
| **Quintile 1** | 9.4% | 11.9% | 2.5% | 8.7% | 3.1% | -5.6% | 13.5% | 4.6% | -8.9% |
| **Quintile 2** | 10.5% | 10.5% | 0.01% | 12.7% | 4.3% | -8.4% | 22.1% | 7.6% | -14.6% |
| **Quintile 3** | 11.0% | 11.9% | 0.9% | 8.4% | 3.6% | -4.8% | 35.4% | 11.7% | -23.7% |
| **Quintile 4** | 13.1% | 13.8% | 0.7% | 10.1% | 4.8% | -5.3% | 29.2% | 9.5% | -19.8% |
| **Quintile 5** | 16.4% | 19.7% | 3.3% | 5.6% | 2.6% | -3.0% | 18.9% | 5.9% | -13.1% |

**Appendix Table 3** – 30-Day Spending Composition of In-Person vs. Telehealth-Initiated Care Episodes for Three Most Common Mental Health and Non-Mental Health CCSR Diagnoses

|  | **Inpatient Spending** | | | **Outpatient Spending** | | | **Part B Spending** | | | **Total Spending** | | |
| --- | --- | --- | --- | --- | --- | --- | --- | --- | --- | --- | --- | --- |
|  | **In-Person** | **Tele**  **health** | **Diff** | **In-Person** | **Tele**  **health** | **Diff** | **In-Person** | **Tele**  **health** | **Diff** | **In-Person** | **Tele**  **health** | **Diff** |
| **Top 3 Mental Health CCSR** | | | | | | | | | | | | |
| **Depression Disorder (MBD002)** | $67 | $18 | -$49 | $17 | $17 | $0 | $186 | $184 | -$2 | $271 | $220 | -$51 |
| **Anxiety and Fear-Related Disorders (MBD005)** | $4 | $1 | -$3 | $10 | $6 | -$4 | $133 | $159 | $26 | $146 | $167 | $20 |
| **Trauma- and Stressor-Related Disorders (MBD007)** | $6 | $1 | -$5 | $6 | $5 | -$1 | $202 | $236 | $34 | $214 | $241 | $28 |
| **Top 3 Non-Mental Health CCSR** | | | | | | | | | | | | |
| **Hypertension (CIR007)** | $0.08 | $0.10 | $0.03 | $11 | $5 | -$6 | $115 | $107 | -$8 | $126 | $112 | -$14 |
| **Sleep Wake Disorders (NVS016)** | $0.17 | $0.09 | -$0.08 | $45 | $18 | -$27 | $110 | $89 | -$21 | $155 | $107 | -$47 |
| **Spondylopathies and Spondyloarthropathy (including infective) (MUS011)** | $161 | $134 | -$27 | $121 | $73 | -$48 | $285 | $208 | -$77 | $568 | $416 | -$152 |

**Appendix Table 4** – 30-Day Return Visit, Imaging, and Lab Test Utilization Rates of In-Person vs. Telehealth-Initiated Care Episodes for Three Most Common Mental Health and Non-Mental Health CCSR Diagnoses

|  | **Return Visit Rate** | | | **Imaging Rate** | | | **Lab Test Rate** | | |
| --- | --- | --- | --- | --- | --- | --- | --- | --- | --- |
|  | **In-Person** | **Telehealth** | **Diff** | **In-Person** | **Telehealth** | **Diff** | **In-Person** | **Telehealth** | **Diff** |
| **Top 3 Mental Health CCSR** | | | | | | | | | |
| **Depression Disorder (MBD002)** | 31.8% | 41.3% | 9.6% | 0.27% | 0.01% | -0.26% | 4.4% | 0.7% | -3.7% |
| **Anxiety and Fear-Related Disorders (MBD005)** | 20.8% | 35.8% | 15.0% | 0.27% | 0.02% | -0.25% | 6.0% | 0.7% | -5.4% |
| **Trauma- and Stressor-Related Disorders (MBD007)** | 41.8% | 59.3% | 17.5% | 0.06% | 0.01% | -0.06% | 3.6% | 0.6% | -2.9% |
| **Top 3 Non-Mental Health CCSR** | | | | | | | | | |
| **Hypertension (CIR007)** | 10.5% | 12.3% | 1.8% | 2.0% | 0.7% | -1.3% | 31.1% | 7.6% | -23.5% |
| **Sleep Wake Disorders (NVS016)** | 6.2% | 7.0% | 0.7% | 0.51% | 0.09% | -0.42% | 11.8% | 5.8% | -6.1% |
| **Spondylopathies and Spondyloarthropathy (including infective) (MUS011)** | 27.8% | 30.3% | 2.5% | 26.6% | 12.5% | -14.1% | 8.0% | 3.8% | -4.2% |

**Appendix Table 5** – 30-Day Spending Composition and Return Visit, Imaging, and Lab Test Utilization Rates of In-Person vs. Telehealth-Initiated Care Episodes Using Matching Without Replacement

|  | **In-Person** | **Telehealth** | **Difference** | **95% Confidence Intervals for Difference** | **P Value** |
| --- | --- | --- | --- | --- | --- |
| **Return Visit Rate** | 14.1% | 16.1% | 2.0% | (2.0% to 2.0%) | <0.001 |
| **Image Rate** | 7.9% | 3.5% | -4.4% | (-4.4% to -4.4%) | <0.001 |
| **Lab Test Rate** | 24.2% | 7.8% | -16.4% | (-16.5% to -16.4%) | <0.001 |
| **Inpatient 30-Day Spending** | $70 | $59 | -$11 | (-$12 to -$10) | <0.001 |
| **Outpatient 30-Day Spending** | $75 | $56 | -$19 | (-$20 to -$19) | <0.001 |
| **Part B 30-Day Spending** | $197 | $145 | -$52 | (-$52 to -$52) | <0.001 |
| **Total 30-Day Spending** | $342 | $260 | -$82 | (-$83 to -$81) | <0.001 |
